# Brain Age: A Promising Biomarker for Understanding Aging in the Context of Cognitive Reserve

**DOI:** 10.1101/2025.01.22.25320988

**Authors:** Iman Beheshti

**Affiliations:** Department of Human Anatomy and Cell Science, Rady Faculty of Health Sciences, University of Manitoba, Winnipeg, MB, Canada

**Keywords:** Aging, Alzheimer’s disease, Brain age prediction, Cognitive decline, Cortical thickness, Hippocampal volume, Longitudinal analysis, Machine learning, T1-weighted MRI

## Abstract

**INTRODUCTION:** Cognitive decline is a major concern in aging populations. Detecting it before clinical symptoms emerge remains a significant challenge. A precise, reliable, and non-invasive biomarker for cognitive health could revolutionize how we monitor normal aging and lifestyle impacts. Such a tool would not only identify individuals at risk of cognitive decline years before symptoms manifest but also aid in early interventions and treatments.

**METHODS:** Longitudinal MRI data from 121 high cognitive reserve (HCR) individuals were compared to matched low cognitive reserve (LCR) individuals to evaluate four biomarkers for early cognitive decline and disease progression: brain age delta, cortical thickness, AD cortical signature, and hippocampal volume. Cross-sectional analyses were conducted at baseline, alongside longitudinal assessments spanning 1 to 12 years, to compare the performance and properties of these biomarkers.

**RESULTS:** The brain age metric emerged as the most reliable biomarker, demonstrating a significant ability to differentiate between groups at baseline (*β* = 1.250, *t* = 3.521, *p* = 0.0009; linear regression model; AUC = 0.73). Furthermore, this biomarker maintained its robustness as the strongest predictor of group membership over a follow-up period of up to 12 years (*β* = 0.409, *p* = 0.025; mixed-effects model), underscoring its potential for longitudinal monitoring of cognitive decline.

**DISCUSSION:** The brain age biomarker demonstrates potential as an effective indicator for early cognitive decline, capable of detecting changes years before clinical symptoms appear and tracking age-related brain and cognitive changes over time. These findings suggest that integrating MRI biomarkers with machine learning approaches could yield more accurate and reliable tools for assessing cognitive health, surpassing the limitations of relying solely on MRI biomarkers.

**Key Points:** Question: Which T1-weighted MRI biomarkers are most effective in predicting longitudinal cognitive deterioration in the aging population?

**Highlights:**

- Four commonly used MRI biomarkers were assessed in the context of cognitive aging.
- Brain age is validated as a promising biomarker for aging and cognitive reserve.
- Machine learning boosts cognitive biomarker accuracy beyond neuroimaging alone.
- The findings underscore a direct association between structural brain reserve and cognitive reserve.
- Key factors in brain preservation may support high cognitive reserve in aging.

## 1. INTRODUCTION

Cognitive decline is a major concern associated with aging, affecting millions of individuals worldwide and presenting significant challenges to healthcare systems and society. As the global population ages, understanding the relationship between aging and cognitive function becomes increasingly important [1, 2]. Aging induces various molecular, cellular, and functional changes in the brain that contribute to cognitive decline, making individuals more vulnerable to neurodegenerative diseases [3]. For instance, genomic instability, telomere dysfunction, and mitochondrial dysfunction in the aging brain lead to chronic oxidative stress, which accelerates aging and age-related diseases, such as cognitive decline [3]. However, not all individuals experience the same trajectory of cognitive decline [4]. Some people maintain cognitive function despite advancing age, a phenomenon referred to as “cognitive reserve” (CR) [4, 5]. CR refers to the brain’s ability to cope with damage and maintain cognitive function, even in the presence of brain pathology, such as Alzheimer’s disease (AD) [4]. Individuals with High Cognitive Reserve (HCR) have greater neural resources and resilience, enabling them to better withstand age-related brain changes or pathology, often maintaining cognitive function longer [6]. In contrast, those with Low Cognitive Reserve (LCR) may experience more pronounced cognitive decline when faced with similar challenges, due to fewer cognitive resources to compensate for neural damage [6]. Factors like education, occupation, and engagement in cognitive activities contribute to the development of CR, enhancing the brain’s resilience to cognitive impairment [4]. Despite growing interest in CR, the underlying mechanisms remain poorly understood, and more research is needed to identify the factors contributing to CR and how they can be leveraged to promote healthier aging [7].

One significant knowledge gap in cognitive aging is the absence of dependable, non-invasive biomarkers that can distinguish individuals with HCR from LCR and many years before symptoms manifest, as well as monitor cognitive function in these groups over time [8, 9]. Timely identification of LCR individuals is essential for implementing interventions that can lessen the effects of neurodegenerative diseases and enhance quality of life [8, 10]. Although clinical assessments can help identify cognitive decline, they often fail to detect changes in the brain until symptoms are already present, making early intervention challenging. As a result, there is increasing reliance on biomarkers, which have the potential to detect subclinical changes in brain structure and function well before cognitive symptoms manifest [11].

Biomarkers for cognitive decline can be classified into several categories, such as fluid biomarkers, neuroimaging biomarkers, and neural activity biomarkers. Fluid biomarkers like amyloid-beta and tau levels in cerebrospinal fluid (CSF) have been used to detect AD before symptoms appear, but they are expensive, invasive, and may not always correlate with cognitive function. Neural activity biomarkers, such as electroencephalography (EEG), can detect abnormal brain activity early, but they lack specificity and do not provide structural information. Neuroimaging biomarkers, particularly MRI, are increasingly used to detect structural changes in the brain, such as hippocampal atrophy and cortical thinning, that can occur well before cognitive symptoms are apparent [12, 13]. One of the primary benefits of neuroimaging biomarkers is the ability for researchers to visually track the impact of aging and/or cognitive decline on brain function and structure.

In recent years, machine learning techniques have been integrated with neuroimaging data to introduce novel biomarkers, such as “brain age” [14, 15]. This method uses neuroimaging data and supervised machine learning algorithms to predict an individual’s “brain age,” providing a quantitative measure of brain health [14, 15]. A deviation from the expected brain age is considered a sign of neurodegeneration or accelerated aging and has shown promise in predicting cognitive decline and disease progression [15, 16].

Among neuroimaging biomarkers, T1-weighted MRI scans are particularly valuable in identifying early signs of cognitive decline and AD. These brain scans can detect structural brain changes and monitor cognitive decline as time passes [12, 13]. Despite the growing body of research on T1-weighted MRI biomarkers in cognitive aging, there remains a significant knowledge gap regarding their ability to identify individuals at risk of cognitive decline years before clinical symptoms appear. Additionally, the effectiveness of these biomarkers in monitoring the progression of cognitive decline over time has been insufficiently explored.

This study aims to address gaps in understanding by evaluating four different T1-weighted MRI biomarkers in the context of cognitive decline. Specifically, we aim to assess their capacity to detect early cognitive decline and monitor brain changes longitudinally. Our focus includes two groups of aging individuals: those who progressed from normal cognition to mild cognitive impairment (MCI) within a 7-year period (LCR group) and those who maintained stable normal cognition over the same period (HCR group). The LCR group may exhibit fewer cognitive resources, contributing to their progression to MCI, whereas the HCR group is hypothesized to possess greater cognitive resources, enabling stable cognition despite aging. Additionally, we will explore the relationship between CR and brain reserve within these groups. The primary objective is to determine which of the four MRI biomarkers most effectively classify individuals into HCR or LCR groups and track cognitive decline over time.

We hypothesize that the brain age biomarker will serve as a robust indicator of early cognitive decline, capable of detecting changes years before clinical symptoms manifest and tracking brain aging and cognitive decline longitudinally.

## 2. MATERIAL AND METHODS

### 2.1. Participants

The dataset for this research was sourced from the Alzheimer’s Disease Sequencing Project Phenotype Harmonization Consortium (ADSP-PHC), a component of the Alzheimer’s Disease Neuroimaging Initiative (ADNI), accessed in September 2024. The ADSP-PHC harmonizes data across multiple cohorts within the ADSP, enabling comprehensive analyses of cognitive functions, neuroimaging, biomarkers, and risk factors. Supported by funding (U24-AG074855), this initiative facilitates advanced genomic research to enhance understanding of AD and Related Dementias (ADRD). Ethical approval was obtained from relevant committees, and all participants provided written informed consent. Detailed methodologies are publicly available via the ADNI database (http://adni.loni.usc.edu/).

The dataset includes over 2,500 individuals with longitudinal T1-weighted MRI scans and clinical variables. Cognitive status was determined using the ‘PHC Diagnosis’ variable and assessed across ADNI1, ADNI2/ADNIGO, and ADNI3 phases. Diagnostic classifications included Cognitively Normal (CN), MCI, and AD. Standard cognitive tests included the Mini-Mental State Examination (MMSE), Clinical Dementia Rating (CDR), Clinical Dementia Rating Scale Sum of Boxes (CDR-SB), and Logical Memory Test.

This study focused on participants cognitively healthy at baseline. A total of 112 participants were classified into the LCR group, defined as transitioning to MCI within a seven-year follow-up, while 134 participants remained cognitively stable, forming the HCR group. In the LCR group, the average transition time to MCI was 3.23 years (±1.32), occurring within 1–5 years. Participants with reversals in cognitive status were excluded.

To balance sample sizes and minimize confounding variables, propensity score matching was applied using the *pymatch* package in Python (https://github.com/benmiroglio/pymatch). A subset of 121 HCR participants was matched to the LCR group based on age, education, sex, MMSE scores, and CDR-SB. Post-matching, both groups exhibited comparable demographic and clinical characteristics, with the matched HCR group serving as an independent test set for analyses.

For brain age estimation model development, additional cognitively healthy samples were incorporated from the ADSP-PHC dataset. The training set consisted of 725 participants (mean age ± SD: 71.56 ± 6.64 years, range: 53–93, 54% female), and the validation set included 81 participants (mean age ± SD: 72.00 ± 7.84 years, range: 52–90, 53% female). All participants had no cognitive or neurological impairments during follow-up. The test dataset of 121 HCR and 121 LCR participants was entirely independent of the training and validation datasets, ensuring the robustness and generalizability of the brain age model.

### 2.2 Image acquisition and processing

T1-weighted MRI data were harmonized across multiple datasets within the Alzheimer’s Disease Sequencing Project (ADSP), encompassing the Alzheimer’s Disease Neuroimaging Initiative (ADNI), National Alzheimer’s Coordinating Center (NACC), Religious Orders Study (ROS), Memory and Aging Project (MAP - Rush), Minority Aging Research Study (MARS), Washington Heights/Inwood Columbia Aging Project (WHICAP), and the Wisconsin Registry for Alzheimer’s Prevention (WRAP).

All T1 scans were preprocessed using a fully automated pipeline with *FreeSurfer* version 6.0 segmentation software (http://surfer.nmr.mgh.harvard.edu). The standardized recon-all cross-sectional *FreeSurfer* pipeline was employed, and quality control (QC) adhered to the ENIGMA QC protocol through visual inspection of surface parcellations. All scans were processed on a consistent computational platform, with QC performed by the same individuals to maintain reliability. To address variations in scanner platforms, models, sites, and field strengths, the Longitudinal COMBAT harmonization technique was applied [17]. The analysis included 68 cortical regions per hemisphere, as defined by the Desikan-Killiany atlas, using volume and thickness values extracted from *aparc.DKTatlas* files [18]. Additionally, 15 subcortical structures were examined, including the thalamus, caudate, putamen, pallidum, hippocampus, and amygdala. Volumetric measures such as total intracranial volume, brain segmentation volumes, and metrics for structures like the lateral ventricles, cerebellum, and brainstem were also included. A total of 197 feature values were extracted for each subject, covering both cortical and subcortical regions. These values were then used as inputs for brain age estimation, generating comprehensive whole-brain data for analysis.

### 2.3 T1-weighted MRI biomarkers

In this study, we concentrated on four established T1-weighted MRI biomarkers: brain age delta, mean cortical thickness, AD cortical signature, and hippocampal volume. These biomarkers are extensively used in research related to neurodegeneration. For each participant, we calculated the mean cortical thickness, which represents the average thickness of the cerebral cortex. The AD cortical signature was derived by assessing the mean cortical thickness in specific regions, including the entorhinal cortex, fusiform gyrus, inferior temporal cortex, and middle temporal cortex [19]. Additionally, the normalized hippocampal volume was determined by adjusting the hippocampal volume relative to the total intracranial volume.

As for the brain age delta, A standard linear support vector regression (SVR) algorithm was employed in MATLAB (i.e., *fitrsvm* function with a linear kernel). In this model, chronological age was the dependent variable, while anatomical measurements extracted from *FreeSurfer* segmentation, along with sex and total intracranial volume, served as independent variables. Initially, we assessed the accuracy of the prediction model on the training dataset through a 10-fold cross-validation strategy. The effectiveness of the model was evaluated using the coefficient of determination (R²), which measures the correlation between chronological age and estimated age, as well as the mean absolute error (MAE) and root mean square error (RMSE). The final brain age estimation model was constructed using the complete training dataset. This model was then applied to independent validation and test sets to calculate the brain age delta, defined as the difference between the predicted brain age and the actual chronological age. To ensure the predicted values were not biased by age, we implemented a bias adjustment technique as outlined in our previous research [20] (https://github.com/Beheshtiiman2/Bias-Correction-in-Brain-Age-Estimation-Frameworks).

### 2.4 Statistical analysis

Student t-test was used to analyze the clinical and demographic differences between the two groups for continuous variables, while the Chi-square test was employed for categorical variables. For baseline data, MRI biomarkers were assessed using a linear regression model to investigate group differences while controlling for age and sex. The model was specified as follows:

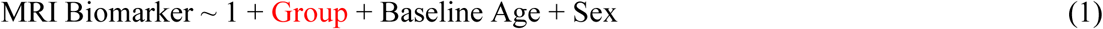

Additional regression models were conducted to examine the effects of sex and age on MRI biomarkers within each group separately. These models were specified as:

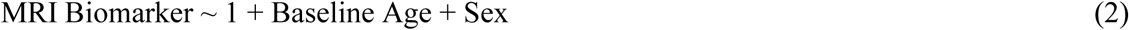

These models were executed independently for each group and combined across groups for baseline data analysis. We further utilized receiver operating characteristic (ROC) curve analysis to compare the accuracy of different MRI biomarkers at baseline.

For the longitudinal follow-up analysis, mixed-effects models were employed to examine cohort differences over time, adjusting for age and sex in each MRI biomarker. The model was specified as:

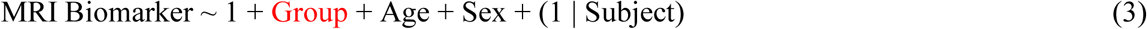

In this mixed-effects model, cohort, age, and sex were treated as fixed effects, while subject was treated as a random effect (Equation 3). To evaluate the effects of age and sex over time within each group, the following mixed-effects model was specified for each MRI biomarker:

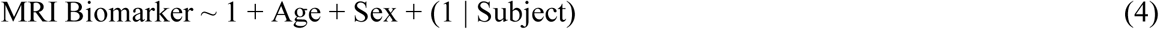

In this model, age, and sex remain fixed effects, with subject continuing to be treated as a random effect. To correct for multiple comparisons, the false discovery rate (FDR) method was used, adjusting the results at a significance level of 0.05. All statistical analyses were conducted using Python.

## 3. RESULTS

### 3.1 Baseline characteristics and clinical features

As intended by the propensity score matching procedure, demographic characteristics and clinical scores at baseline were not significantly different between the HCR and LCR individuals (Table 1). There were no significant differences in the percentage of females, chronological age, years of education, MMSE scores, or CT (*p* > 0.05 for all). However, LCR participants had slightly higher CDR-SB scores compared to HCR participants (*p* = 0.0431). A significant difference was observed in the proportion of APOE4 carriers between the LCR group (68/37/6) and the HCR group (84/27/1) (*p* = 0.0395). After false discovery rate (FDR) correction, the two groups were similar in all demographic and clinical features (*p* > 0.22).

**Table 1.** Demographic and Clinical Features of the Test Sets at Baseline.

|  | HRC<br>(N=112) | LCR<br>(N=112) | Original P-Value | Corrected P-Value |
| --- | --- | --- | --- | --- |
| Female (%) | 50% | 42% | 0.1805 | 0.2707 |
| Real age (yrs) | 74.02 ± 4.8 | 75.00 ± 5.9 | 0.1725 | 0.2707 |
| Education (yrs) | 16.42±2.8 | 16.34±2.6 | 0.8250 | 0.8250 |
| MMSE | 29.02±1.21 | 28.90±1.27 | 0.3622 | 0.4346 |
| CDR-SB | 0.03±0.64 | 0.06±0.93 | 0.0431 | 0.1293 |
| APOE4 (0/1/2) | 84/27/1 | 68/37/6 | 0.0395 | 0.1293 |

Note: CDR-SB: Clinical Dementia Rating Sum of Boxes; LCR: Low Cognitive Reserve; HRC: High Cognitive Reserve; MMSE: Mini-Mental State Examination. Mean ± standard deviation is presented for all variables. Original P-Value: Results from Student's t-test for continuous variables and Chi-square test for categorical variables between groups. Corrected P-Value: Original p-value adjusted for FDR correction.

### 3.2 Brain age estimation performance

Our predictive model demonstrated robust performance on the training dataset (N = 725) with 10-fold cross-validation, yielding an R² of 0.85, an MAE of 2.10 years, and an RMSE of 2.68 years (mean brain age delta = 0 ± 3.8 years, Fig. 1). The model’s performance on the validation set (N = 81) was consistent, with an R² of 0.90, an MAE of 2.07 years, an RMSE of 2.63 years, and a mean brain age delta of 0.055 ± 3.8 years. The correlation between brain age delta and chronological age was non-significant in both the training set (r = 0, *p* = 1) and the validation set (r = 0.03, *p* = 0.77).

**Figure 1:**
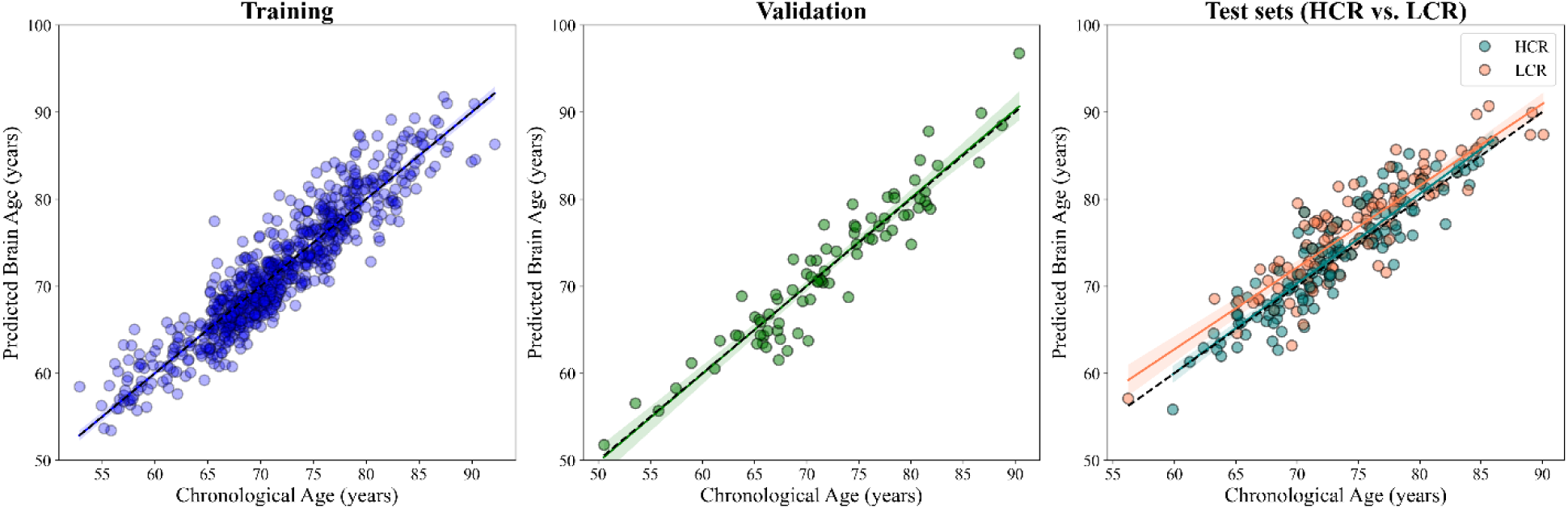
Scatter plots showing the association between predicted brain age and chronological age across the Training, Validation, and Test datasets. The results for the Training set were generated through 10-fold cross-validation. The dashed black line in all panels represents the identity line (y=x) for reference.

### 3.3 Baseline MRI Metric Analysis

At baseline, participants in the LCR group exhibited significantly lower normalized hippocampal volume (*p* = 0.0011) and a higher brain age delta (*p* = 0.0006), suggesting an accelerated aging process in this cohort. Additionally, the AD cortical signature was more pronounced in the LCR group (*p* = 0.0039). In contrast, both the LCR and HCR groups displayed similar mean cortical thickness at baseline (*p* = 0.1786). After applying FDR correction, the differences in the AD cortical signature (*p* = 0.0130), normalized hippocampal volume (*p* = 0.0055), and brain age delta (*p* = 0.0055) remained statistically significant between the two groups (Table 2 and Fig. 2).To further evaluate the discriminative ability of various brain measures, we conducted a ROC analysis to differentiate between the LCR and HCR groups at baseline (Fig. 3). The brain age delta demonstrated the highest discriminatory power with an Area Under the Curve (AUC) of 0.73. In comparison, hippocampal volume (AUC = 0.63), AD cortical signature (AUC = 0.60), and mean cortical thickness (AUC = 0.55), showed limited accuracy in distinguishing between the groups.

**Table 2.** Baseline MRI biomarkers for independent test groups.

|  | HRC<br>(N=112) | LCR<br>(N=112) | Original P-Value | Corrected P-Value |
| --- | --- | --- | --- | --- |
| Mean cortical thickness (mm) | 2.32±0.09 | 2.30±0.12 | 0.1786 | 0.2256 |
| AD Cortical signature (mm) | 2.87±0.13 | 2.81±0.16 | 0.0039 | 0.0130 |
| Normalized hippocampal volume (%) | 0.50 ±0.5 | 0.47 ± 0.6 | 0.0011 | 0.0055 |
| Brain age delta (yrs) | 0.40±2.68 | 1.84±25.91 | 0.0006 | 0.0055 |

Note: LCR: Low Cognitive Reserve; HRC: High Cognitive Reserve; Mean ± standard deviation is presented for all variables. Original P-Value: Results from Student's t-test for continuous variables and Chi-square test for categorical variables between groups. Corrected P-Value: Original p-value adjusted for FDR correction.

**Figure 2:**
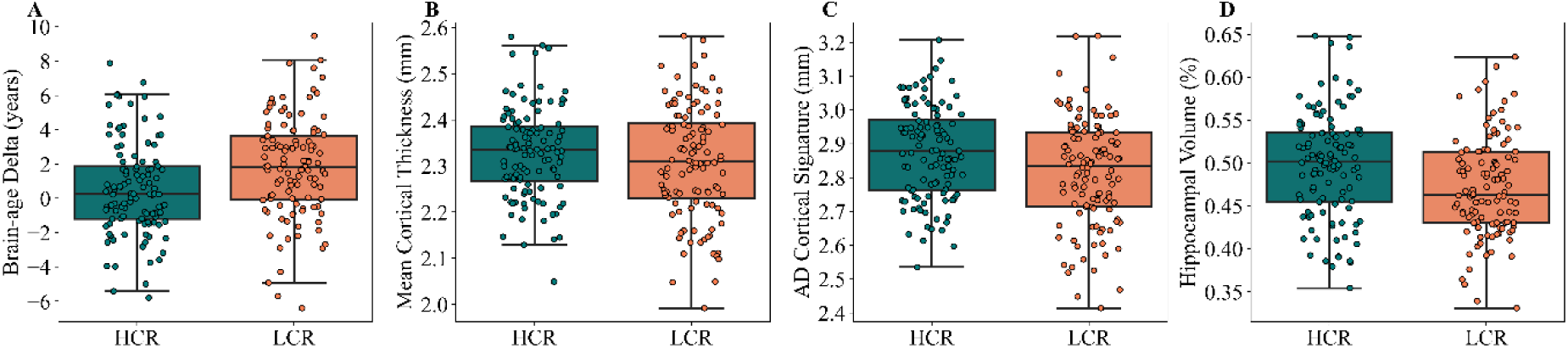
Box plots depicting brain age delta (years), mean cortical thickness (mm), AD cortical signature (mm), and hippocampal volume (%) for the LCR and HRC groups at the baseline. Statistically significant differences were observed between the two groups for brain age delta, AD cortical signature, and hippocampal volume at baseline. No significant difference was found for mean cortical thickness.

**Figure 3:**
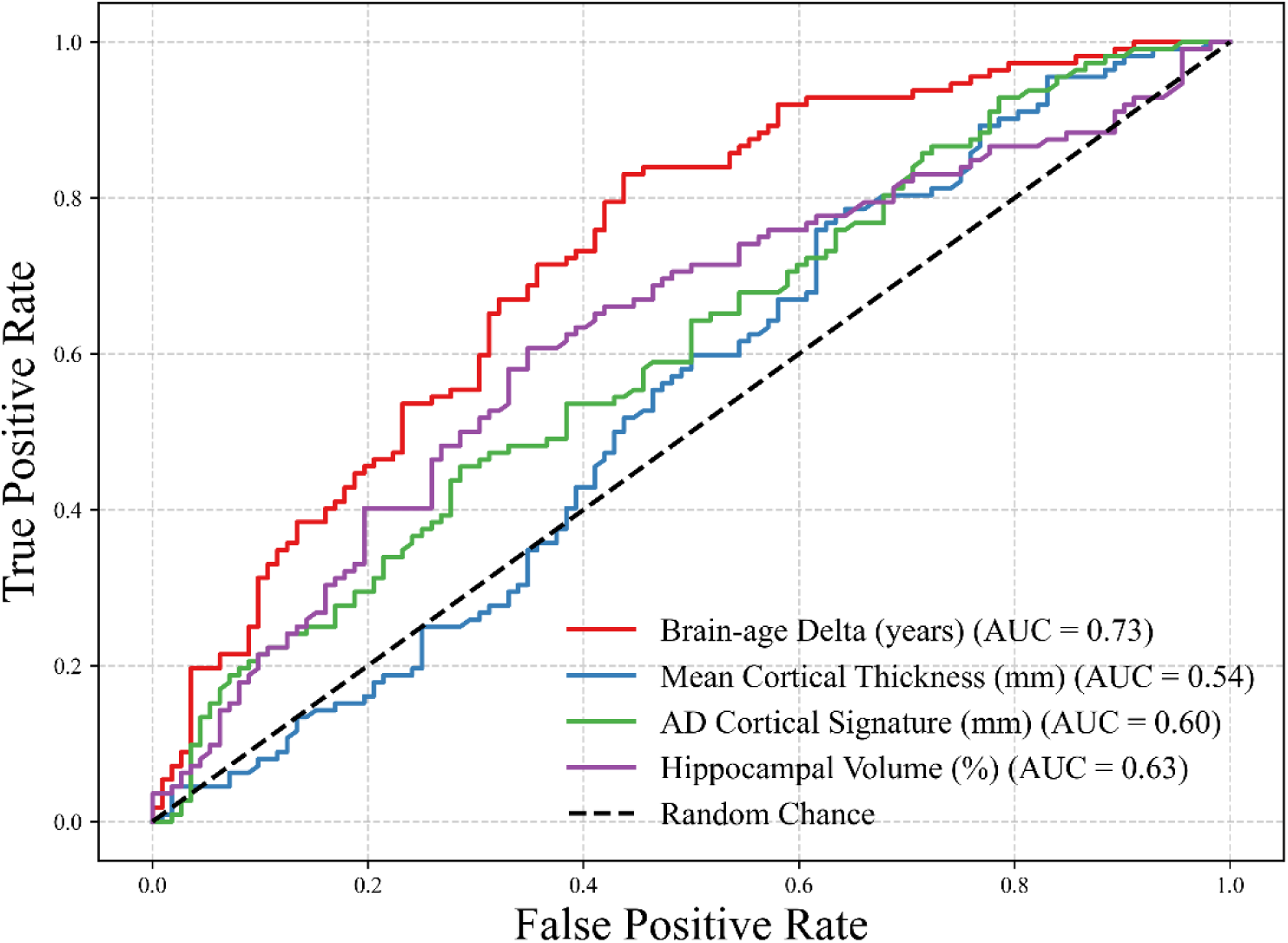
Receiver Operating Characteristic (ROC) curves comparing the ability of four MRI-derived measures to differentiate between the LCR and HRC groups at baseline.

Linear regression analysis was conducted to assess the relationship between MRI biomarkers, group status, age, and sex (Table 3). Brain age delta positively associated with group status (*β* = 1.250, *p* = 0.0009) and significantly correlated with sex (*β* = -0.948, p = 0.0118), indicating accelerated brain aging in the LCR group and sex-related differences. Mean cortical thickness did not differ between groups (*β* = 0.0091, *p* = 0.5085) but correlated with age (*β* = - 0.006, *p* < 0.0001) and sex (*β* = 0.0473, *p* = 0.00077). The AD cortical signature showed lower values in LCR (*β* = -0.0502, *p* = 0.0103) and correlated significantly with age (*β* = -0.0078, *p* < 0.0001). Normalized hippocampal volume was lower in the LCR group (*β* = -0.0206, *p* = 0.0063), with significant associations with age (*β* = -0.0033, *p* = 0.00098) and sex (*β* = 0.0259, *p* = 0.0006) (Table 3).

**Table 3:** Regression model outputs on different MRI metrices at baseline.

| Model |  | Brain Age Delta (years) | Mean Cortical Thickness (mm) | AD Cortical Signature (mm) | Hippocampal Volume (%) |
| --- | --- | --- | --- | --- | --- |
| Group | $\beta$ | 1.250 | 0.0091 | -0.0502 | -0.0206 |
|  | t | 3.521 | -0.6622 | -2.5848 | 2.757 |
|  | p | 0.0009* | 0.5085 | 0.0103 | 0.0063* |
| Actual age | $\beta$ | -0.042 | -0.006 | -0.0078 | -0.0033 |
|  | t | -1.229 | -4.883 | -4.317 | -4.737 |
|  | p | 0.221 | 0.000002* | 0.00002* | 0.00098 |
| Sex | $\beta$ | -0.948 | 0.0473 | 0.00379 | 0.0259 |
|  | t | -2.536 | 3.4091 | 0.19499 | 3.441 |
|  | p | 0.0118 | 0.00077* | 0.8455 | 0.0006* |

Note: An asterisk (\*) denotes p-values that remained statistically significant after FDR p-value correction.

At baseline, there was no significant correlation between brain age delta and age for either group (Fig. 4). However, when stratified by sex, only the LCR group demonstrated a significant difference between males and females, with males exhibiting higher brain age delta (t = 3.78, *p* = 0.008; Fig. 5). As expected, mean cortical thickness, AD cortical signature, and hippocampal volume followed a clear age-related pattern, with older age associated with lower values (Fig. 4, Table 3). In the HCR group, males had significantly lower hippocampal volume compared to females, though no significant differences between sexes were observed for other metrics (Table 4). In the LCR group, in addition to brain age delta, mean cortical thickness also differed significantly between sexes, with males showing lower values (t = 3.89, *p* = 0.001; Fig. 5B, Table 4).

**Figure 4:**
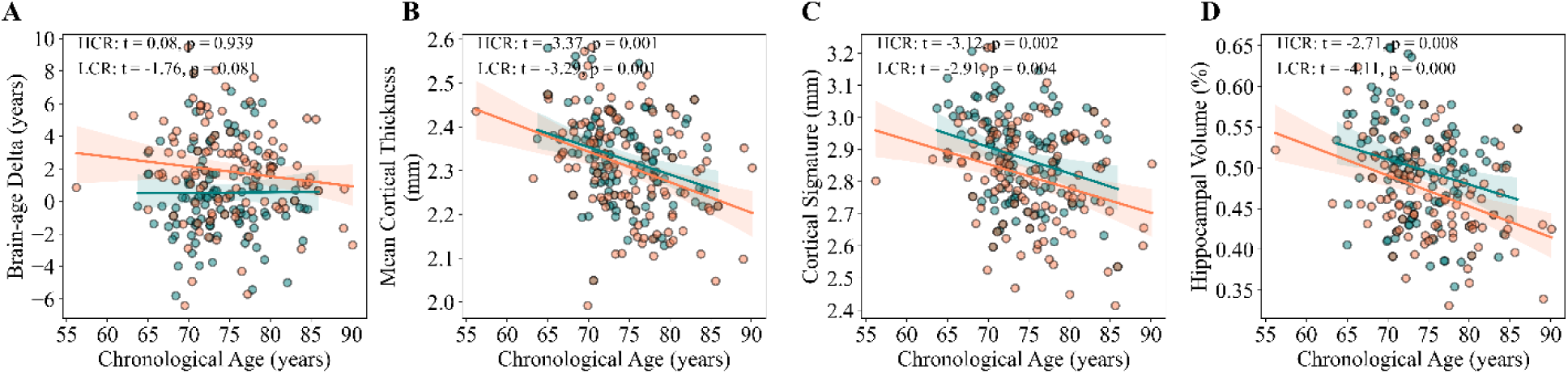
Association between different MRI biomarkers and age in HCR and LCR groups.

**Figure 5:**
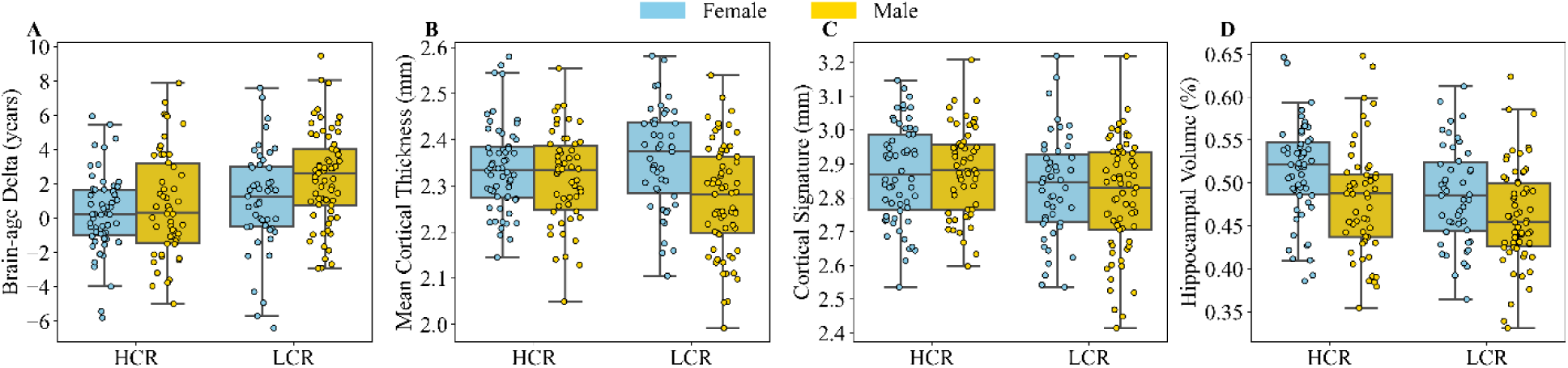
Boxplots illustrating the distribution of different MRI biomarkers in the HCR and LCR groups according to sex.

**Table 4:** Regression Model Results for Baseline MRI Metrics and Associations with Age and Sex.

| Group | Model |  | Brain Age Delta (years) | Mean Cortical Thickness (mm) | AD Cortical Signature (mm) | Hippocampal Volume (%) |
| --- | --- | --- | --- | --- | --- | --- |
| HCR | Actual age | $\beta$ | 0.004 | -0.006 | -0.008 | -0.003 |
|  |  | t | 0.077 | -3.365 | -3.118 | -2.710 |
|  |  | p | 0.939 | 0.001* | 0.002* | 0.008* |
| | Sex | $\beta$ | -0.506 | 0.021 | -0.005 | 0.034 |
|  |  | t | -0.992 | 1.201 | -0.201 | 3.133 |
|  |  | p | 0.324 | 0.232 | 0.841 | 0.002* |
| LCR | Actual age | $\beta$ | -0.081 | -0.006 | -0.007 | -0.004 |
|  |  | t | -1.760 | -3.291 | -2.910 | -4.109 |
|  |  | p | 0.081 | 0.001* | 0.004* | 0.000* |
| | Sex | $\beta$ | -1.482 | 0.075 | 0.014 | 0.016 |
|  |  | t | -2.700 | 3.489 | 0.454 | 1.601 |
|  |  | p | 0.008* | 0.001* | 0.651 | 0.112 |

Note: An asterisk (\*) denotes p-values that remained statistically significant after FDR p-value correction.

### 3.4 Longitudinal analysis

The LCR and HCR groups exhibited distinct trends over time in terms of MMSE and CDR-SB scores. Specifically, HCR subjects showed relatively stable patterns in both MMSE and CDR-SB, while these clinical cognitive assessments deteriorated over time in the LCR group (Fig. 6). Differences between the two groups across MRI metrics over a 12-year follow-up, along with the results of the generalized linear mixed-effects model examining these group differences, are presented in Fig. 7 and Table 5. The strongest differentiator was brain age delta, where group membership showed a significant effect (*β* = 0.409, *p* = 0.025). This was followed by the AD cortical signature, which also revealed a significant difference between groups (*β* = -0.024, *p* = 0.020). In contrast, hippocampal volume exhibited a marginal trend toward significance (β = - 0.005, *p* = 0.094), suggesting potential group differences. Mean cortical thickness did not show any significant group effect (β = -0.010, *p* = 0.167), indicating no distinguishing power between the groups for this measure. Demographic factors, such as sex and age, were significant predictors across many metrics, with sex influencing brain age delta, cortical thickness, and hippocampal volume, while age at baseline consistently had a negative impact on cortical thickness, hippocampal volume, and AD cortical signature.

**Figure 6:**
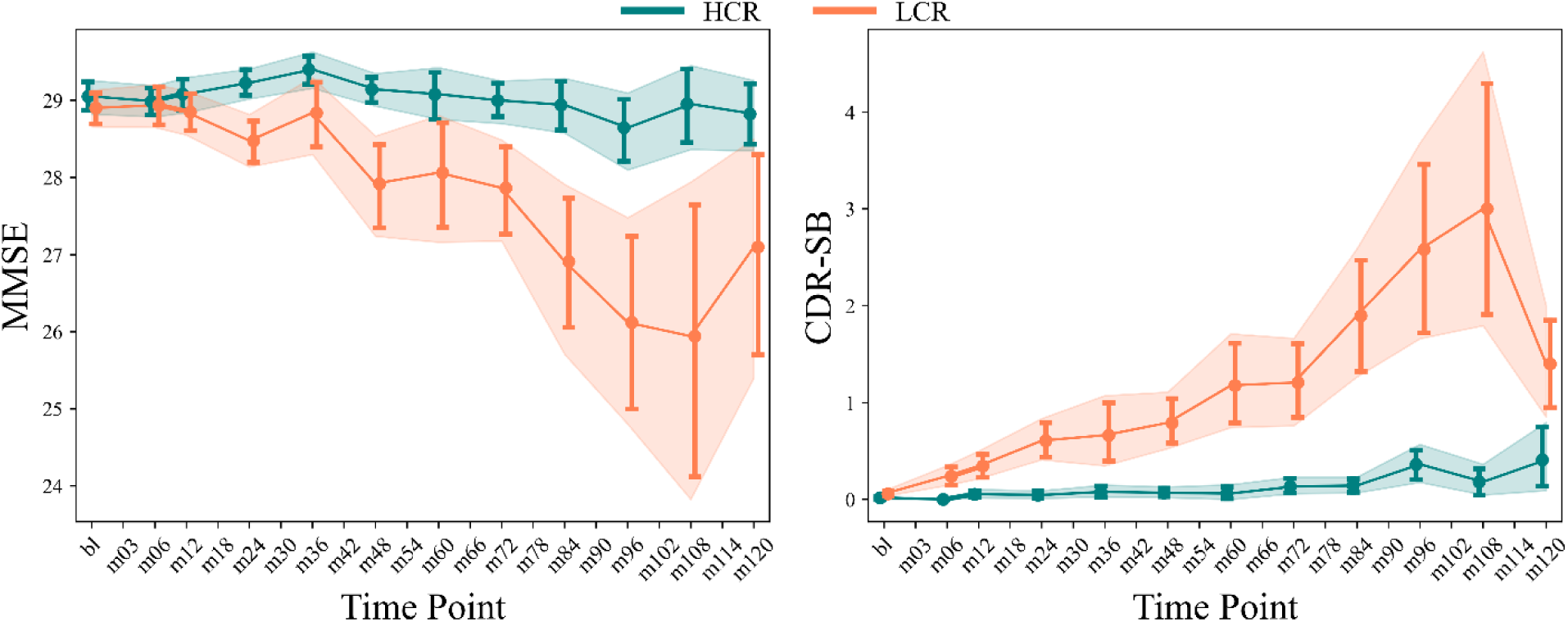
Point plot showing the pattern of MMSE and CDR-SB two groups over a 12-year follow-up. Markers (’o’) indicate mean values at various time points, with error bars representing the 90% confidence interval (CI). “bl” denotes baseline data, and “mXX” indicates months post-baseline (e.g., 03, 06, 12, …, 120 months).

**Figure 7:**
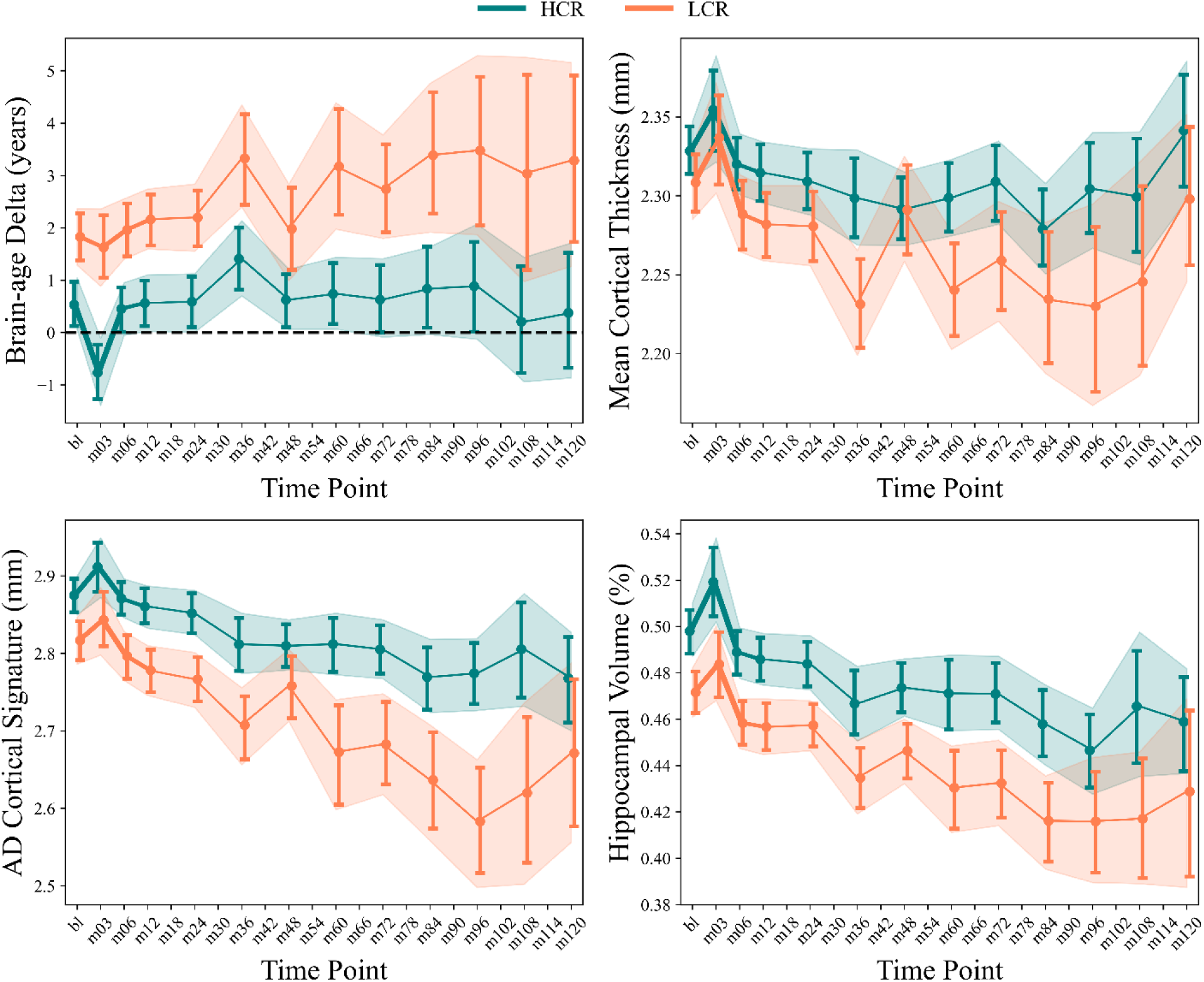
Point plot showing differences between two groups across MRI metrics over a 12-year follow-up. Markers (’o’) indicate mean values at various time points, with error bars representing the 90% confidence interval (CI). “bl” denotes baseline data, and “mXX” indicates months post-baseline (e.g., 03, 06, 12, …, 120 months).

**Table 5:** Results of the generalized linear mixed-effects model examining group differences across MRI metrics.

| | $\beta$ | S.E. | z | P | 95% CI |
| --- | --- | --- | --- | --- | --- |
| **Brain Age Delta** |  |  |  |  |  |
| Intercept | 2.163 | 1.006 | 2.150 | 0.032* | [0.191, 4.135] |
| Group | 0.409 | 0.183 | 2.237 | 0.025* | [0.051, 0.768] |
| Sex | -0.909 | 0.395 | -2.301 | 0.021 | [-1.684, -0.135] |
| Age | 0.002 | 0.012 | 0.148 | 0.882 | [-0.021, 0.025] |
| 1 \| Subject | -0.000 | 0.000 | -3.607 | 0.000* | [-0.001, 0.000] |
| **Mean Cortical Thickness** |  |  |  |  |  |
| Intercept | 2.439 | 0.039 | 63.031 | 0.000* | [2.363, 2.515] |
| Group | -0.010 | 0.007 | -1.381 | 0.167 | [-0.023, 0.004] |
| Sex | 0.044 | 0.013 | 3.323 | 0.001* | [0.018, 0.070] |
| Age | -0.002 | 0.000 | -5.388 | 0.000* | [-0.003, -0.002] |
| 1 \| Subject | 0.000 | 0.000 | 4.985 | 0.000* | [0.000, 0.000] |
| **AD Cortical Signature** |  |  |  |  |  |
| Intercept | 3.751 | 0.057 | 65.934 | 0.000* | [3.639, 3.862] |
| Group | -0.024 | 0.010 | -2.321 | 0.020* | [-0.044, -0.004] |
| Sex | -0.006 | 0.020 | -0.306 | 0.760 | [-0.045, 0.033] |
| Age | -0.012 | 0.001 | -18.114 | 0.000* | [-0.014, -0.011] |
| 1 \| Subject | 0.000 | 0.000 | 1.718 | 0.086 | [-0.000, 0.000] |
| **Hippocampal Volume** |  |  |  |  |  |
| Intercept | 0.766 | 0.016 | 47.688 | 0.000* | [0.734, 0.797] |
| Group | -0.005 | 0.003 | -1.674 | 0.094 | [-0.010, 0.001] |
| Sex | 0.018 | 0.007 | 2.576 | 0.010* | [0.004, 0.032] |
| Age | -0.004 | 0.000 | -22.221 | 0.000* | [-0.004, -0.004] |
| 1 \| Subject | 0.000 | 0.000 | 3.598 | 0.000* | [0.000, 0.000] |

Note: S.E. = Standard Error. The models were based on Equation 3 and included a total of 1,585 observations across 242 samples. The "Group" variable was the primary variable of interest in this analysis. An asterisk (\*) denotes p-values that remained statistically significant after FDR p-value correction.

A linear mixed model was employed to investigate the impact of age and sex on each MRI metric, considering the random effects of participants (Table 6). In both groups, sex did not significantly impact brain age delta. However, age had differing effects in each group: the brain age delta decreased with age in the HCR group (*β* = -0.053, *p* < 0.001), while it increased with age in the LCR group (*β* = 0.095, *p* < 0.001). Regarding mean cortical thickness, a significant sex difference was found in the LCR group, with females exhibiting a relatively faster or more positive change compared to males. The age-related decline in cortical thickness was more pronounced in the LCR group (*β* = -0.005, *p* < 0.001) than in the HCR group (*β* = -0.001, *p* = 0.085), suggesting a stronger effect of age on cortical thickness for LCR individuals. For the AD cortical signature, no significant sex effects were observed in either group. In both groups, significant negative associations between age and AD cortical signature were noted, with the LCR group exhibiting a stronger effect (*β* = -0.018, *p* < 0.001) compared to the HCR group (*β* = -0.009, *p* < 0.001). Lastly, a sex difference was observed in the HCR group for hippocampal volume, where males showed a relatively faster or more positive change compared to females. The impact of age on hippocampal volume was significant in both groups, showing a more pronounced negative correlation in the LCR group (*β* = -0.005, *p* < 0.001) than in the HCR group (*β* = -0.003, *p* < 0.001).

**Table 6:** Results from the generalized linear mixed-effects model assessing age and sex differences in MRI metrics across each group.

| | Group | $\beta$ | S.E. | z | P | 95% CI |
| --- | --- | --- | --- | --- | --- | --- |
| **Brain Age Delta** |  |  |  |  |  |  |
| Intercept | HCR | 5.888 | 1.152 | 5.113 | 0.000* | [3.631, 8.145] |
|  | LCR | -4.089 | 1.768 | -2.312 | 0.021* | [-7.555, -0.623] |
| Sex | HCR | -0.507 | 0.467 | -1.086 | 0.277 | [-1.422, 0.408] |
|  | LCR | -0.728 | 0.614 | -1.186 | 0.236 | [-1.930, 0.475] |
| Age | HCR | -0.053 | 0.014 | -3.901 | 0.000* | [-0.080, -0.026] |
|  | LCR | 0.095 | 0.021 | 4.507 | 0.000* | [0.053, 0.136] |
| 1 \| Subject | HCR | -0.000 | 0.000 | -4.175 | 0.000* | [-0.001, -0.000] |
|  | LCR | -0.000 | 0.000 | -1.914 | 0.056 | [-0.001, -0.000] |
| **Mean Cortical Thickness** |  |  |  |  |  |  |
| Intercept | HCR | 2.352 | 0.047 | 49.651 | 0.000* | [2.259, 2.445] |
|  | LCR | 2.576 | 0.064 | 40.477 | 0.000* | [2.451, 2.700] |
| Sex | HCR | 0.024 | 0.017 | 1.424 | 0.155 | [-0.009, 0.058] |
|  | LCR | 0.061 | 0.020 | 3.117 | 0.002* | [0.023, 0.099] |
| Age | HCR | -0.001 | 0.001 | -1.722 | 0.085 | [-0.002, 0.000] |
|  | LCR | -0.005 | 0.001 | -6.207 | 0.000* | [-0.006, -0.003] |
| 1 \| Subject | HCR | 0.000 | 0.000 | 2.894 | 0.004* | [0.000, 0.000] |
|  | LCR | 0.000 | 0.000 | 4.276 | 0.000* | [0.000, 0.000] |
| **AD Cortical Signature** |  |  |  |  |  |  |
| Intercept | HCR | 3.499 | 0.064 | 54.665 | 0.000* | [3.373, 3.624] |
|  | LCR | 4.146 | 0.102 | 40.492 | 0.000* | [3.945, 4.346] |
| Sex | HCR | -0.021 | 0.023 | -0.918 | 0.358 | [-0.066, 0.024] |
|  | LCR | -0.018 | 0.033 | -0.536 | 0.592 | [-0.082, 0.047] |
| Age | HCR | -0.009 | 0.001 | -11.433 | 0.000* | [-0.010, -0.007] |
|  | LCR | -0.018 | 0.001 | -14.711 | 0.000* | [-0.020, -0.016] |
| 1 \| Subject | HCR | 0.000 | 0.000 | 2.656 | 0.008* | [0.000, 0.000] |
|  | LCR | 0.000 | 0.000 | 0.821 | 0.412 | [-0.000, 0.000] |
| **Hippocampal Volume** |  |  |  |  |  |  |
| Intercept | HCR | 0.722 | 0.021 | 34.297 | 0.000* | [0.681, 0.763] |
|  | LCR | 0.832 | 0.024 | 34.496 | 0.000* | [0.785, 0.879] |
| Sex | HCR | 0.025 | 0.010 | 2.484 | 0.013* | [0.005, 0.044] |
|  | LCR | 0.005 | 0.009 | 0.505 | 0.614 | [-0.013, 0.023] |
| Age | HCR | -0.003 | 0.000 | -14.356 | 0.000* | [-0.004, -0.003] |
|  | LCR | -0.005 | 0.000 | -18.169 | 0.000* | [-0.006, -0.005] |
| 1 \| Subject | HCR | 0.000 | 0.000 | 2.767 | 0.006* | [0.000, 0.000] |
|  | LCR | 0.000 | 0.000 | 3.314 | 0.001* | [0.000, 0.000] |

Note: S.E. = Standard Error. The models were based on Equation 4 and conducted separately for each group. Age and sex were the primary variables of interest in this analysis. An asterisk (\*) denotes p-values that remained statistically significant after FDR p-value correction.

## 4. DISCUSSION

The primary aim of this study was to evaluate the reliability of various T1-weighted MRI biomarkers in distinguishing between HCR and LCR in aging individuals, well before clinical symptoms manifest. We focused on four validated biomarkers: brain age delta, mean cortical thickness, AD cortical signature, and hippocampal volume, all of which are widely utilized in neurodegeneration research. The brain age delta estimates biological brain age using structural MRI features, revealing a brain metric that correlates with cognitive outcomes [14–16]. A higher brain age delta is linked to poorer cognitive performance and is associated with neurodegenerative disorders like AD, showing significant correlations with clinical measures such as MMSE and CDR [21]. Cortical thickness measures the thickness of the cerebral cortex and is indicative of cognitive function. Cortical thinning is associated with CSF protein levels linked to regulatory, developmental, and inflammatory processes, which play a key role in age-related cortical thinning [22]. Thinning is often observed in AD, particularly in memory-related regions, making it a valuable tool for differentiating AD from normal aging. However, the relationship between cortical structure and AD remains complex. The AD Cortical Signature represents a specific pattern of cortical thinning in AD, particularly in the medial temporal lobe and frontal areas [19]. This signature is crucial for early detection and monitoring of AD progression and has shown superior predictive utility in distinguishing MCI from dementia. Hippocampal Volume is a key biomarker for memory and spatial navigation, with reductions indicating AD. Maintaining hippocampal volume is associated with better cognitive health in aging, and it serves as a reliable indicator of disease progression in AD research. Regarding brain age delta, our prediction model demonstrated a desirable MAE of < 2.10 years on both training and validation sets. This performance aligns well with similar studies that used T1-weighted data for constructing brain age estimation frameworks [23–25].

### 4.1 Cross-Sectional Analysis

Regarding brain age delta, a significant positive coefficient was observed, indicating that individuals with LCR exhibited a higher brain age delta compared to HCR (*β* = 1.250, t = 3.521, *p* = 0.0009). This suggests that LCR individuals experience accelerated brain aging, even when their cognitive function remains healthy. This finding aligns with research indicating that brain age delta is a sensitive biomarker for neurodegenerative processes and cognitive decline [26]. The strong association between group membership and brain age delta highlights its potential as a valuable tool for identifying individuals at higher risk for neurological disorders, even in the absence of clinical diagnoses [26]. Interestingly, no significant difference in mean cortical thickness was observed between the two groups at the baseline (*β* = 0.0091, t = -0.6622, *p* = 0.5085). The absence of a significant difference here may suggest that global cortical thinning is not a distinguishing feature between aging with LCR and aging with HCR groups, particularly when the two groups are matched in terms of age, sex, and education. Alternatively, localized changes (e.g., AD cortical signature) was relevant in this context. The significant difference in AD cortical signature between groups (*β* = -0.0502, *p* = 0.0103) reveals distinct patterns of cortical atrophy or structural changes that differentiate aging with LCR and aging with HCR. This biomarker offers insight into specific brain regions affected by AD and could play a key role in early detection and monitoring of neurodegenerative processes. In examining hippocampal volume, a significant negative coefficient was found at baseline (*β* = -0.0206, t = 2.757, *p* = 0.0063), indicating that individuals with LCR properties have lower hippocampal volume compared to those with HCR properties. This suggests that LCR individuals may be at greater risk for memory-related issues and cognitive decline. Since the hippocampus is crucial for memory formation, its reduced volume in those with LCR properties could reflect underlying neurological changes that may not yet be clinically evident.

Based on statistical significance and effect sizes, brain age delta demonstrated the strongest group effect (highest t-value, lowest p-value), followed by hippocampal volume. Although AD cortical signature showed a significant group effect, it was less pronounced than brain age delta and hippocampal volume. The ROC analysis supported these findings, showing that brain age delta had the highest discriminatory power (AUC = 0.73), while cortical thickness (AUC = 0.55), AD cortical signature (AUC = 0.60), and hippocampal volume (AUC = 0.63) showed limited accuracy. Our results surpass those of previous studies, which reported an AUC of 0.656 for distinguishing cognitively healthy aging individuals who later converted to MCI from those who did not, based on chronological age, sex, and brain age delta derived from structural MRI data [26].Unlike our study, the mentioned study did not match groups for baseline age, sex distribution, or MMSE scores, and significant differences were observed in other baseline demographic and clinical variables [26].

### 4.2 Longitudinal analysis

In our longitudinal analysis, we noticed that aging with HCR and aging with LCR exhibit distinct patterns as time progresses. The results of the generalized linear mixed-effects model reveal significant insights into the differences in MRI metrics between groups (Table 5). Notably, brain age delta (*p* = 0.025) and AD Cortical Signature (*p* =0.020) emerged as the most effective measures for distinguishing between LCR and HCR groups, with both showing statistically significant differences. The positive coefficient for brain age delta suggests that LCR group exhibits an older brain age compared to the other over time, indicating potential neurodegenerative changes. Similarly, the negative coefficient for the AD Cortical Signature implies that LCR group may have higher cortical thickness shrinking in the specific cortical thickness regions. In contrast, mean cortical thickness (*p* = 0.167) and hippocampal volume (*p* = 0.094) did not demonstrate significant differences, suggesting they may be less reliable indicators in this context. Overall, these findings highlight the ability of brain age delta to trace cognitive decline over time in aging with HCR and aging with LCR. This highlights the potential of brain age biomarkers for use in clinical assessments and research on neurodegenerative diseases.

Interesting results were observed when assessing the impact of age on each group separately using linear mixed models (Table 6). Age had a significant negative effect on brain age delta in the HCR group (*β* = -0.053, *p* < 0.001), meaning that as individuals age, their brain age delta decreases, suggesting better cognitive resilience. In contrast, the LCR group showed a significant positive effect of age (*β* = 0.095, *p* < 0.001), indicating that older individuals in this group have a greater brain age delta, reflecting accelerated aging. In other biomarkers, we observed a negative association with age. As for the cortical thickness biomarker, age negatively impacted mean cortical thickness in both groups, with the LCR group showing a more pronounced effect (*β* = -0.005, p < 0.001) compared to the HCR group (*β* = -0.001, *p* = 0.085). This indicates that cortical thinning is more significant in the LCR group as they age. This pattern was also observed on AD cortical thickness biomarker such age had a significant negative effect on the AD cortical signature for both groups (HCR: *β* = -0.009, *p* < 0.001; LCR: *β* = -0.018, *p* < 0.001), indicating that older individuals exhibit a more pronounced AD cortical signature, which is consistent with the progression of Alzheimer’s pathology, and LCR group showed faster AD cortical signature shrinking than HCR. As for hippocampal volume, age negatively affected in both groups (HCR: *β* = -0.003, p < 0.001; LCR: *β* = -0.005, p < 0.001), indicating that as individuals age, hippocampal volume decreases, which is a common finding in neurodegenerative conditions, but this event occur a bit faster in LCR group. In general, in this analyse, brain age delta showed a robust and excellent results as it showed opposite trends in HCR and LCR groups, making it the most effective biomarker for distinguishing between the two groups over time than other T1-weighted MRI biomarkers. Previous studies have identified an association between accelerated cognitive decline and increased rates of brain volume loss [27]. In our LCR group, we observed a strong relationship between disease progression (from cognitive health to MCI) and brain characteristics, specifically cortical thickness and hippocampal volumes. This suggests that faster rates of cognitive decline are associated not only with more rapid brain volume loss but also with localized cortical thinning, such as the AD cortical signature.

Our longitudinal findings also underscore a robust connection between structural brain preservation and cognitive resilience. Specifically, in our HCR group, age-related brain atrophy and cortical thinning occurred at a significantly slower pace compared to LCR group. This observation highlights the critical role of protective factors in mitigating substantial brain atrophy. Key factors contributing to this preservation may include genetics [28, 29], physical activity [30, 31], life satisfaction [30], dietary patterns [31, 32], diabetes [30], sleep quality [33], social engagement, education [34], and environmental influences [35]. Despite these insights, the precise molecular mechanisms by which these factors shield the brain from significant atrophy remain unclear [35]. Further research is essential to unravel these underlying processes, which could inform strategies for enhancing cognitive and structural brain health in aging populations. These results not only emphasize the importance of fostering these protective factors throughout life but also offer valuable direction for future studies aimed at improving the quality of life and cognitive function among older adults.

### 4.3 Impact of age and sex factors

An interesting observation was the lack of significant correlation between brain age delta and chronological age when we examined group differences in cross-sectional and longitudinal analyses (Tables 3 and 5). This result can be attributed to the use of a validated bias adjustment technique, which minimizes the influence of age on brain-age values [20]. A major advantage of a non-age-dependent biomarker is its broad applicability, eliminating the need for age-based adjustments. Such biomarkers provide more consistent disease indications, making them useful across different age groups and avoiding complications from normal age-related changes. In contrast, mean cortical thickness, AD cortical signature, and hippocampal volume showed strong age-related patterns, with older age correlating with lower values in both cross-sectional and longitudinal analyses (Tables 3 and 5). This age dependence complicates the interpretation and generalizability of results, as age must be considered a confounding factor. It has been reported that cognitively healthy males tend to have brains appearing significantly older than their female counterparts, with an average difference of 5.58 years [36]. This tendency has also been observed in terms of glucose brain metabolism, with cognitively healthy males showing a significantly higher mean metabolic brain age difference compared to cognitively healthy females, with a difference of 1.1 years [37]. Consistent with these findings, our analysis revealed that cognitively healthy males exhibited a higher brain age delta compared to females at baseline when data from both groups were combined (N = 242; t = 2.73, *p* = .006, independent t-test; mean difference = 1 year). When comparing aging with HCR and LCR, a significant sex difference was observed only in the LCR group. Specifically, males in the LCR group exhibited a notably higher brain age delta (t = 3.78, p = .008; Fig. 5A) and a significantly lower mean cortical thickness than females (t = 3.89, *p* = .001, independent t-test; Fig. 5B, Table 4). In the group with HCR, males had significantly lower hippocampal volume than females, although no significant differences were observed between sexes for other metrics (Table 4). These findings suggest that sex may play a complex role in cognitive decline and the progression from cognitively healthy states to MCI or AD. Further research is needed to better understand the impact of sex on these transitions.

To the best of our knowledge, only a small number of studies have explored the progression from cognitively healthy to MCI in elderly individuals, with most relying primarily on baseline data [26, 38, 39]. Moreover, detecting the onset of MCI from a cognitively healthy state remains a significant challenge due to the subtle nature of morphological and cognitive changes in the early stages [40–42]. While some studies have investigated this transition, our study provides novel insights by leveraging a variety of MRI biomarkers to address this complex process. What sets our research apart from previous studies is its unique methodological strengths. Our analysis utilized both cross-sectional and longitudinal data, following participants for a minimum of 7 years, with some cases extending up to 12 years. This extended follow-up allowed us to categorize participants into HCR and LCR groups, enabling a deeper understanding of how each group behaves over time in relation to MRI biomarkers. Unlike prior studies that classified participants based solely on cognitive status, our approach included neuroimaging validation of MCI diagnosis. To ensure the highest quality data, we employed pre-processed MRI features using validated software such as *FreeSurfer*, renowned for its robustness in multi-center and multi-scanner studies [43]. Additionally, to mitigate potential confounding effects from varying scanner manufacturers, we applied a harmonization technique, longitudinal *ComBat*, to standardize the MRI features [17]. Our study also benefited from the harmonized data provided by the ADSP-PHC, further enhancing the reliability of our results. A key strength of our analysis was the inclusion of data from multiple sites and scanners (Section 2), which underscores the generalizability of our findings. The predictive model used in our study demonstrated strong performance even when tested on independent datasets, further validating its robustness. Importantly, we ensured that the groups were matched for baseline characteristics, including age, sex distribution, and cognitive scores (MMSE, CDR, and CDR-SB). This careful matching ensures that the observed structural brain differences between the groups can be attributed to neuroimaging features, rather than baseline demographic or cognitive variations. In general, our cross-sectional and longitudinal analyses highlight that brain age delta offers more robust results compared to other T1-weighted MRI biomarkers for distinguishing LCR aging individuals from HCR aging individuals, even years before clinical symptoms appear, when both groups exhibit normal cognitive functioning. This finding emphasizes the potential of combining machine learning algorithms with MRI data to improve clinical outcomes, rather than relying solely on MRI data. Our study’s comprehensive approach, combining rigorous methodology with a robust dataset, provides new insights into the brain’s structural changes associated with cognitive decline. Despite the strength of our findings, it is important to note that additional studies utilizing other neuroimaging modalities, such as tau and amyloid positron emission tomography (PET), diffusion tensor imaging (DTI), and functional MRI, are needed. These studies would not only help validate our results but also provide new insights into the underlying mechanisms of cognitive reserve in the aging population.

## 5. Conclusion

While further research is required to determine the optimal T1-weighted MRI-based biomarker for detecting LCR aging, our findings suggest that brain age holds great promise as a key biomarker for understanding brain structural aging. It has the potential to identify individuals at risk for MCI long before clinical symptoms appear, as well as to monitor cognitive performance and assess the effectiveness of interventions and treatments over time. Moreover, our study demonstrates that integrating machine learning algorithms with neuroimaging data enhances the accuracy of cognitive biomarkers, underscoring the transformative role of machine learning in advancing health sciences. Additionally, our findings highlight the direct connection between structural brain reserve and cognitive reserve, demonstrating that individuals with HCR exhibited significantly less structural brain change with increasing age. This underscores the importance of protective factors (e.g., physical activity, life satisfaction, dietary patterns, diabetes management, sleep quality, social engagement, education, and environmental influences) in promoting both brain reserve and cognitive reserve.

## Data Availability

Data, Ethical agreements and Code Availability
The data used in this study were obtained from ADNI database, as outlined in Section 2. Ethical approval was secured for the ADNI project, and all participants provided written informed consent. Access to ADNI data is available to researchers through a Data Use Agreement (DUA), which ensures adherence to ethical standards, including the protection of participant confidentiality and proper data handling.

## ACKNOWLEDGMENTS

We would like to thank the principal investigators of the Alzheimer’s Disease Neuroimaging Initiative (ADNI) dataset for providing the data used in this article. Their efforts in collecting and making these datasets accessible are greatly appreciated. We also want to express our gratitude to the participants who volunteered to be part of these datasets. Their cooperation and participation have been crucial in furthering our knowledge in this area.

- ADNI which was funded by National Institutes of Health (Grant U01 AG024904) and DOD ADNI (Department of Defense award number W81XWH-12-2-0012), the National Institute on Aging, the National Institute of Biomedical Imaging and Bioengineering, and through generous contributions from the following: AbbVie, Alzheimer’s Association; Alzheimer’s Drug Discovery Foundation; Araclon Biotech; BioClinica, Inc.; Biogen; Bristol-Myers Squibb Company; CereSpir, Inc.; Cogstate; Eisai Inc.; Elan Pharmaceuticals, Inc.; Eli Lilly and Company; EuroImmun; F. Hoffmann-La Roche Ltd and its affiliated company Genentech, Inc.; Fujirebio; GE Healthcare; IXICO Ltd.; Janssen Alzheimer Immunotherapy Research & Development, LLC.; Johnson & Johnson Pharmaceutical Research & Development LLC.; Lumosity; Lundbeck; Merck & Co., Inc.; Meso Scale Diagnostics, LLC.; NeuroRx Research; Neurotrack Technologies; Novartis Pharmaceuticals Corporation; Pfizer Inc.; Piramal Imaging; Servier; Takeda Pharmaceutical Company; and Transition Therapeutics. The Canadian Institutes of Health Research is providing funds to support ADNI clinical sites in Canada. Private sector contributions are facilitated by the Foundation for the National Institutes of Health (www.fnih.org). The grantee organization is the Northern California Institute for Research and Education, and the study is coordinated by the Alzheimer’s Therapeutic Research Institute at the University of Southern California. ADNI data are disseminated by the Laboratory for Neuro Imaging at the University of Southern California.

## Data, Ethical agreements and Code Availability

The data used in this study were obtained from ADNI database, as outlined in Section 2. Ethical approval was secured for the ADNI project, and all participants provided written informed consent. Access to ADNI data is available to researchers through a Data Use Agreement (DUA), which ensures adherence to ethical standards, including the protection of participant confidentiality and proper data handling. Brain age estimation and bias correction were conducted using a framework that has been previously validated and is available for access at https://github.com/Beheshtiiman2/Bias-Correction-in-Brain-Age-Estimation-Frameworks.

## REFERENCES

1. Deary, I.J., et al., Age-associated cognitive decline. British medical bulletin, 2009. 92(1): p. 135–152.

2. Jessen, F., et al., The characterisation of subjective cognitive decline. The Lancet Neurology, 2020. 19(3): p. 271–278.

3. Cordeiro, A., et al., Aging and cognitive resilience: Molecular mechanisms as new potential therapeutic targets. Drug Discovery Today, 2024: p. 104093.

4. Nelson, M.E., et al., Cognitive reserve, Alzheimer’s neuropathology, and risk of dementia: A systematic review and meta-analysis. Neuropsychology review, 2021. 31(2): p. 233–250.

5. Pettigrew, C. and A. Soldan, Defining cognitive reserve and implications for cognitive aging. Current neurology and neuroscience reports, 2019. 19: p. 1–12.

6. Stern, Y., What is cognitive reserve? Theory and research application of the reserve concept. Journal of the international neuropsychological society, 2002. 8(3): p. 448–460.

7. Stern, Y., et al., Whitepaper: Defining and investigating cognitive reserve, brain reserve, and brain maintenance. Alzheimer’s & Dementia, 2020. 16(9): p. 1305–1311.

8. Counts, S.E., et al., Biomarkers for the early detection and progression of Alzheimer’s disease. Neurotherapeutics, 2017. 14(1): p. 35–53.

9. De Looze, C., et al., Assessing cognitive function in longitudinal studies of ageing worldwide: some practical considerations. Age and ageing, 2023. 52(Supplement_4): p. iv13–iv25.

10. Whelan, R., et al., Developments in scalable strategies for detecting early markers of cognitive decline. Translational Psychiatry, 2022. 12(1): p. 473.

11. Hansson, O., Biomarkers for neurodegenerative diseases. Nature medicine, 2021. 27(6): p. 954–963.

12. Chouliaras, L. and J.T. O’Brien, The use of neuroimaging techniques in the early and differential diagnosis of dementia. Molecular Psychiatry, 2023. 28(10): p. 4084–4097.

13. Sun, J., J.-D.J. Han, and W. Chen, Exploring the relationship among Alzheimer’s disease, aging and cognitive scores through neuroimaging-based approach. Scientific Reports, 2024. 14(1): p. 27472.

14. Sone, D. and I. Beheshti, Neuroimaging-based brain age estimation: A promising personalized biomarker in neuropsychiatry. Journal of Personalized Medicine, 2022. 12(11): p. 1850.

15. Mishra, S., I. Beheshti, and P. Khanna, A review of neuroimaging-driven brain age estimation for identification of brain disorders and health conditions. IEEE Reviews in Biomedical Engineering, 2021. 16: p. 371–385.

16. Gaser, C., P. Kalc, and J.H. Cole, A perspective on brain-age estimation and its clinical promise. Nature computational science, 2024: p. 1–8.

17. Beer, J.C., et al., Longitudinal ComBat: A method for harmonizing longitudinal multi-scanner imaging data. Neuroimage, 2020. 220: p. 117129.

18. Klein, A. and J. Tourville, 101 labeled brain images and a consistent human cortical labeling protocol. Front Neurosci, 2012. 6: p. 171.

19. Jack Jr, C.R., et al., Different definitions of neurodegeneration produce similar amyloid/neurodegeneration biomarker group findings. Brain, 2015. 138(12): p. 3747–3759.

20. Beheshti, I., et al., Bias-adjustment in neuroimaging-based brain age frameworks: A robust scheme. NeuroImage: Clinical, 2019. 24: p. 102063.

21. Beheshti, I., N. Maikusa, and H. Matsuda, The association between “Brain-Age Score”(BAS) and traditional neuropsychological screening tools in Alzheimer’s disease. Brain and Behavior, 2018. 8(8): p. e01020.

22. Ekblad, L.L., et al., Proteomic correlates of cortical thickness in cognitively normal individuals with normal and abnormal cerebrospinal fluid beta-amyloid1-42. Neurobiology of aging, 2021. 107: p. 42–52.

23. Jónsson, B.A., et al., Brain age prediction using deep learning uncovers associated sequence variants. Nature communications, 2019. 10(1): p. 5409.

24. Baecker, L., et al., Machine learning for brain age prediction: Introduction to methods and clinical applications. EBioMedicine, 2021. 72.

25. Dörfel, R.P., et al., Prediction of brain age using structural magnetic resonance imaging: A comparison of accuracy and test–retest reliability of publicly available software packages. Human Brain Mapping, 2023. 44(17): p. 6139–6148.

26. Choi, U.-S., et al., Predicting mild cognitive impairments from cognitively normal brains using a novel brain age estimation model based on structural magnetic resonance imaging. Cerebral Cortex, 2023. 33(21): p. 10858–10866.

27. Armstrong, N.M., et al., Associations between cognitive and brain volume changes in cognitively normal older adults. Neuroimage, 2020. 223: p. 117289.

28. Kaufmann, T., et al., Common brain disorders are associated with heritable patterns of apparent aging of the brain. Nature neuroscience, 2019. 22(10): p. 1617–1623.

29. Brouwer, R.M., et al., Genetic variants associated with longitudinal changes in brain structure across the lifespan. Nature neuroscience, 2022. 25(4): p. 421–432.

30. Sone, D., et al., Neuroimaging-derived brain age is associated with life satisfaction in cognitively unimpaired elderly: A community-based study. Translational psychiatry, 2022. 12(1): p. 25.

31. Dominguez, L.J., et al., Nutrition, physical activity, and other lifestyle factors in the prevention of cognitive decline and dementia. Nutrients, 2021. 13(11): p. 4080.

32. Arora, S., et al., Diet and lifestyle impact the development and progression of Alzheimer’s dementia. Frontiers in Nutrition, 2023. 10: p. 1213223.

33. Sehar, U., et al., Effects of sleep deprivation on brain atrophy in individuals with mild cognitive impairment and Alzheimer’s disease. Ageing research reviews, 2024: p. 102397.

34. Steffener, J. and Y. Stern, Exploring the neural basis of cognitive reserve in aging. Biochimica et Biophysica Acta (BBA)-Molecular Basis of Disease, 2012. 1822(3): p. 467–473.

35. Yang, Z., et al., Brain aging patterns in a large and diverse cohort of 49,482 individuals. Nature medicine, 2024. 30(10): p. 3015–3026.

36. Cole, J.H., et al., Brain age predicts mortality. Molecular psychiatry, 2018. 23(5): p. 1385–1392.

37. Beheshti, I., et al., Disappearing metabolic youthfulness in the cognitively impaired female brain. Neurobiology of aging, 2021. 101: p. 224–229.

38. Wee, C.-Y., et al., Cortical graph neural network for AD and MCI diagnosis and transfer learning across populations. NeuroImage: Clinical, 2019. 23: p. 101929.

39. Mofrad, S.A., et al., Cognitive and MRI trajectories for prediction of Alzheimer’s disease. Scientific reports, 2021. 11(1): p. 2122.

40. Roberts, R.O., et al., Higher risk of progression to dementia in mild cognitive impairment cases who revert to normal. Neurology, 2014. 82(4): p. 317–325.

41. Lipnicki, D.M., et al., Risk factors for mild cognitive impairment, dementia and mortality: the Sydney memory and ageing study. Journal of the American Medical Directors Association, 2017. 18(5): p. 388–395.

42. Toledo, J.B., et al., Neuronal injury biomarkers and prognosis in ADNI subjects with normal cognition. Acta neuropathologica communications, 2014. 2: p. 1–9.

43. Dadar, M., S. Duchesne, and C. Group, Reliability assessment of tissue classification algorithms for multi-center and multi-scanner data. NeuroImage, 2020. 217: p. 116928.

